# Xpert MTB/RIF^®^ cycle threshold as a marker of TB disease severity; Implications for TB treatment stratification

**DOI:** 10.1101/2025.06.12.25329126

**Authors:** Daniel J. Grint, Jasvir Dhillon, Philip D. Butcher, Jack Adams, Tulika Munshi, Adam A. Witney, Katherine Gould, Kenneth Laing, Christopher Cousins, Sean Wasserman, Katherine Fielding, Tom Harrison, Amina Jindani, the RIFASHORT study team

**Affiliations:** Department of Infectious Disease Epidemiology, London School of Hygiene and Tropical Medicine, London, WC1E 7HT; Institute for Infection and Immunity, City St George’s, University of London, London; Wellcome Discovery Research Platforms in Infection, Centre for Infectious Diseases Research in Africa, Institute of Infectious Disease and Molecular Medicine, University of Cape Town, Observatory, Cape Town, Republic of South Africa

**Author notes:** Corresponding author: Daniel Grint, London School of Hygiene and Tropical Medicine.

**Keywords:** Sputum culture, sputum smear, chest X-ray, TB phenotype, relapse, Ultra

## Abstract

**Introduction:** Recent trials have demonstrated that shortened four-month treatment durations are effective for the majority of people with tuberculosis (TB). However, there is a population of TB patients who require longer treatment durations. Prospectively identifying those who require shorter versus longer treatment durations would support evaluation and implementation of optimized regimens.

**Methods:** We analysed data from the RIFASHORT TB treatment-shortening non-inferiority trial to define a TB phenotype classification. The RIFASHORT trial primary outcome was reanalysed using the protocol-defined non-inferiority criterion of eight percentage points, stratifying by those classified as having limited or extensive disease.

**Results:** Xpert MTB/RIF® semiquantitative bacterial burden in combination with TB disease involvement grading on chest X-ray achieved the strongest differentiation between relapse and non-relapse. The extensive disease TB phenotype (high semiquantitative bacterial burden and extensive TB disease on X-ray) accounted for one quarter of the RIFASHORT trial population and more than half of all post-treatment TB relapses (13/23). For the limited TB disease phenotype (<high semiquantitative bacterial burden or no extensive TB disease on X-ray), the experimental 4-month 1200mg rifampicin-containing regimen met the protocol-defined non-inferiority criterion in both modified-intention-to-treat (adjusted risk difference: -1.3% (95% confidence interval: -6.7% to 4.0%)) and per protocol analyses (1.7% (95% CI: -3.8% to 7.1%)).

**Conclusion:** A four-month treatment duration, with a widely available, tolerable, rifampicin-based TB treatment regimen, was non-inferior to the standard of care for three-quarters of people with TB in the RIFASHORT trial. This finding justifies definitive evaluation of disease-stratified rifampicin-based TB treatment in a phase III randomised trial.

**summary:** Xpert MTB/RIF® and chest radiography at TB treatment initiation identify a majority of patients for whom a 4-month treatment duration is non-inferior to the standard of care. These widely available measures may facilitate personalised TB treatment durations.

## Introduction

Studies of drug-susceptible TB treatment regimens shorter than 6-months have demonstrated that, for a large proportion of people, treatment duration can be reduced to less than 6-months with no adverse impact on outcomes.^1-4^ However, all but one of these trials failed to show non-inferiority of the shorter regimens in the overall study population. The question remains how to identify people with TB disease who may be adequately treated with shorter treatment durations as opposed to those who require a longer duration of treatment.

Established measures of TB disease severity, such as sputum smear grade, and abnormalities on chest X-ray are known to be associated with TB treatment outcomes, but are poor prognostic tools for individual responses.^5^ Previous attempts to individualise TB treatment duration according to sputum smear and culture status have proven unsuccessful.^6,7^

Cepheid Xpert® MTB/RIF and Xpert® MTB/RIF Ultra are two widely-used diagnostic tools for TB disease.^8,9^ These assays output quantitative cycle threshold values and a semiquantitative assessment (high, medium, low, very low, trace) of bacterial burden, which are being explored as correlates of disease severity.^10,11^

Xpert® MTB/RIF cycle threshold presents a near-patient measure of TB disease severity that is easily obtained and interpreted with results available within two hours. An accessible test which accurately differentiates TB disease severity and poor treatment outcome risk is highly desirable and may enable individualised TB treatment durations.

This study assessed Xpert® MTB/RIF and Xpert® MTB/RIF Ultra cycle threshold as a measure of TB disease severity in comparison with other established measures. We defined a TB phenotype classification of limited and extensive disease based on baseline measures of TB disease severity. In addition, we reanalysed data from a recently completed phase III trial of four-month TB treatment regimens, stratifying the analysis by the TB phenotype classification.

## Methods

### Study population and data

We analysed data from the RIFASHORT trial, which assessed two 4-month TB treatment regimens both containing an increased dose of rifampicin (1200mg or 1800mg) provided once-daily as a flat dose together with standard doses of isoniazid, plus ethambutol and pyrazinamide for 2 months, compared with the 6-month standard of care containing rifampicin at 10 mg/kg (300 mg to 750 mg once-daily, according to weight band). Participants were enrolled in Botswana, Guinea, Nepal, Pakistan, Peru, and Uganda with the primary outcome defined at 12-months post-randomisation. The trial composite primary unfavourable outcome included culture confirmed TB relapse, retreatment for TB, death or loss to follow-up during treatment, and permanent treatment change due to adverse events. Details on the study design are available in the main trial results publication.^3^ In brief, 672 HIV-negative adults with pulmonary TB were randomised 1:1:1 to the three study arms. Neither experimental regimen demonstrated non-inferiority in comparison to the control, based on a non-inferiority margin of eight percentage points. However, a high proportion of participants had a successful treatment outcome in all three trial arms (Control: 93%; 1200mg rifampicin: 90%; 1800mg rifampicin: 87%).

Baseline data were collected on sputum smear grade, sputum culture grade (defined below), chest X-ray parameters, and cycle threshold from either Xpert® MTB/RIF (Xpert) or Xpert® MTB/RIF Ultra (Ultra), depending on which test platform was available at each site. The Xpert and Ultra platforms differ with respect to their gene targets. Xpert has five probes targeting sequences of the *rpoB* gene, whereas Ultra includes an additional gene signature (IS1081/IS6110) as well as four probes targeting *rpoB* gene sequences.^8,9^ For both tests, a quantitative cycle threshold output is derived from the *rpoB* gene probe outputs. A lower cycle threshold, the number of cycles required to detect TB, is indicative of a higher bacterial burden. Both tests also output a semiquantitative interpretation of the cycle threshold value, indicating whether the bacterial burden is high, medium, low, very low, or trace (the latter for Ultra only). Interpretation of the semiquantitative bacterial burden is consistent between Xpert and Ultra as the quantitative cycle threshold cut-points defining the semiquantitative bacterial burden was calibrated independently for each test.^8,9^

Baseline chest X-rays were collated digitally and read centrally by an independent expert at St George’s University of London. All sputum cultures were performed on solid LJ-slope medium at local site laboratories. Recurrence samples were stored locally at least -20 degrees centigrade and shipped to St George’s University of London for whole genome sequencing.

### Statistical Methods

The study population included all RIFASHORT trial participants who tested positive on Xpert or Ultra and had baseline cycle threshold data. Post-treatment TB relapse was defined by two consecutive positive sputum cultures with no evidence of exogenous reinfection by whole genome sequencing. Positive last available sputum cultures with TB treatment reinitiated, but without a confirmatory culture, were also defined as TB relapses.

#### Xpert® MTB/RIF cycle threshold as a measure of TB disease severity

The distribution of Xpert and Ultra cycle threshold probe values at baseline were described using boxplots, pooling data from the whole study population.

Quantitative cycle threshold values were computed as the minimum of the *rpoB* probe outputs. The association between Xpert and Ultra cycle threshold quantitative output and other measures of disease severity (sputum smear, sputum culture, and chest X-ray) was described using boxplots and assessed formally using linear regression. For sputum smear, the Ziehl-Neelsen grading system was used (no acid-fast bacilli seen, scanty, +, ++, +++). Sputum cultures were graded according to the number of bacterial colonies seen on LJ slopes (+: 20-100; ++: 100-200; +++: >200).

The likelihood ratio test assessed for general association between each measure of disease severity and Xpert or Ultra cycle threshold quantitative output. The coefficient of determination (R^2^) was used to describe the proportion of variability in Xpert and Ultra cycle threshold quantitative output explained by other disease severity markers. To understand if there were inherent differences in the populations tested with Xpert and Ultra, we also compared TB disease severity markers between these two groups.

#### TB phenotype classification

In determining optimal markers for classifying limited and extensive TB disease, data were pooled from the two four-month study arms. The control arm was excluded to avoid treatment efficacy biasing this analysis as there were fewer TB relapses in the control arm. The sensitivity and specificity of each disease severity marker in identifying TB relapse was estimated along with two-way combinations of each disease severity marker combined with Xpert and Ultra semiquantitative bacterial burden.

For chest X-ray we explored three grading systems. The first used a study-specific classification system graded by an independent expert as: far advanced disease, moderate disease, minimal disease, or normal (defined in supplement Section 1). The second grading combined three chest X-ray observations (≥50% lung involvement, cavitation, and bilateral disease) into a binary classification. The third grading was a binary classification based on lung involvement alone (≥50%).

#### Reanalysis of RIFASHORT trial data by TB phenotype classification

Reanalysis of RIFASHORT trial data followed the same methods as the primary analysis of the trial, including modified intention-to-treat and per-protocol analyses (further details in supplement Section 2). We evaluated the risk difference in the proportion with an unfavourable treatment outcome at 12-months post-randomisation using Cochran-Mantel-Haenszel weights and the protocol-defined non-inferiority criterion of eight percentage points, with stratification by study site. The analysis was performed separately for those classified as having limited and extensive TB disease.

## Results

### Relapse characteristics

99.3% (667/672) of RIFASHORT trial participants were Xpert or Ultra positive and included in the analysis. Baseline TB disease severity measures of the study population are shown in Table 1. Ultra was used for 21% of participants in Peru, 37% of participants in Uganda, and 59% of participants in Guinea. Xpert was used exclusively in Botswana, Nepal, or Pakistan. A high semiquantitative bacterial burden accounted for 30.3% of participants tested with Xpert and 62.6% of participants tested with Ultra.

**Table 1.** Population characteristics.

| Population Characteristics | N (%)<br>(N=667) | Relapse (row %)<br>(N=25) |
| --- | --- | --- |
| Study site |  |  |
| Botswana | 54 (8.1) | 2 (3.7) |
| Peru | 119 (17.8) | 0 |
| Uganda | 224 (33.6) | 15 (6.7) |
| Nepal | 70 (10.5) | 3 (4.3) |
| Guinea | 175 (26.2) | 5 (2.9) |
| Pakistan | 25 (3.7) | 0 |
| Sex |  |  |
| Male | 490 (73.5) | 22 (4.5) |
| Female | 177 (26.5) | 3 (1.7) |
| Missing | 8 (1.2) | 0 |
| Age in years; Median (IQR) | 29 (23 - 39) | 38 (24 - 46) |
| BMI; Median (IQR) | 18.7 (17.0 – 20.9) | 18.1 (16.3 – 19.9) |
| Chest X-ray extent of lung disease |  |  |
| Normal | 2 (0.3) | 0 |
| Minimal disease | 18 (2.7) | 0 |
| Moderately advanced disease | 294 (44.1) | 7 (2.4) |
| Far advanced | 343 (51.4) | 18 (5.2) |
| Missing | 4 (0.6) | 0 |
| Cavitation on chest X-ray |  |  |
| Yes | 578 (86.7) | 23 (4.0) |
| No | 85 (12.7) | 2 (2.4) |
| Bilateral disease on chest X-ray |  |  |
| Yes | 446 (66.9) | 22 (3.3) |
| No | 219 (32.8) | 3 (0.5) |
| Missing | 2 (0.3) | 0 |
| Chest X-ray lung zone involvement score |  |  |
| 0 | 6 (0.9) | 0 |
| 1 | 28 (4.2) | 0 |
| 2 | 119 (17.8) | 1 (0.8) |
| 3 | 171 (25.6) | 6 (3.5) |
| 4 | 176 (26.4) | 9 (5.1) |
| 5 | 98 (14.7) | 7 (7.1) |
| 6 | 65 (9.7) | 2 (3.1) |
| Extensive disease on chest X-ray* |  |  |
| Yes | 274 (41.1) | 4 (1.5) |
| No | 389 (58.3) | 21 (5.4) |
| Missing | 4 (0.6) | 0 |
| Baseline sputum smear grade |  |  |
| No AFB seen | 45 (6.7) | 1 (2.2) |
| Scanty / + | 250 (37.5) | 4 (1.6) |
| ++ | 148 (22.2) | 11 (7.4) |
| +++ | 224 (33.6) | 9 (4.0) |
| Missing | 7 (1.0) | 0 |
| Baseline sputum culture grade |  |  |
| Negative | 27 (4.0) | 2 (7.4) |
| + | 140 (21.0) | 3 (2.1) |
| ++ | 160 (24.0) | 5 (3.1) |
| +++ | 333 (49.9) | 15 (4.5) |
| Baseline Xpert MTB/RIF semi-quantitative CT (N=456) |  |  |
| High | 138 (30.3) | 8 (5.8) |
| Medium | 218 (47.8) | 6 (2.8) |
| Low | 76 (16.7) | 2 (2.6) |
| V. Low | 24 (5.3) | 0 |
| Baseline Xpert MTB/RIF Ultra semi-quantitative CT (N=211) |  |  |
| High | 132 (62.6) | 8 (6.1) |
| Medium | 60 (28.4) | 1 (1.7) |
| Low | 14 (6.6) | 0 |
| V. Low | 7 (3.3) | 0 |

AFB: acid-fast bacilli; CT: cycle threshold.
\*Extensive disease on chest X-ray was defined as $\geq 50\%$ lung involvement, bilateral disease, and cavitation.

There were 25 TB relapses during post-treatment follow-up, 22 of which were culture confirmed, giving an overall relapse risk of 3.7% (25/667). Two relapses occurred in the control arm, with 11 and 12 in the 1200mg and 1800mg rifampicin treatment arms, respectively.

There was no difference in the distribution of chest X-ray lung grading scores between participants tested with Xpert or Ultra (P=0.16), nor lung involvement zone scores (P=0.34), or sputum smear grading (P=0.13). However, a higher proportion of those tested with Ultra had cavitation on chest X-ray (91.5% vs. 84.4%; P=0.03) and had 3+ on sputum culture (56.8% vs. 46.7%; P<0.0001), compared with those tested with Xpert.

The risk of TB relapse during follow-up was highest in Uganda (6.7%) and Nepal (4.3%). There were no relapses among participants in Peru and Pakistan. Men had a higher relapse risk than woman (4.5% vs. 1.7%) and relapse cases were older (median: 38 vs. 28 years). There were no TB relapses among people with normal or minimal disease on chest X-ray, although these groups were small (n=20 for both combined). Relapse risk was 2.4% and 5.2% among those with moderately advanced disease and far advanced disease on chest X-ray, respectively. According to sputum smear, the highest relapse risk was seen for those with a 2+ grading (7.4%). For sputum culture, the highest relapse risk was seen for those with a +++ grading (4.5%). For Xpert and Ultra semiquantitative bacterial burden, the highest relapse risk was in the high grading (5.8% and 6.1%, respectfully).

### Xpert MTB/RIF and Ultra cycle threshold and disease severity

There was higher variability in the distribution of quantitative cycle threshold values from Xpert compared to Ultra (Figure 1). Ultra *rpoB* probe cycle threshold values were strongly right-skewed Ultra IS probe cycle threshold values had very low variability, clustering around the median value (median: 16.4 (interquartile range 16.1 – 17.0)), emphasising that the IS probe is not designed to be interpreted in terms of disease burden.

**Figure 1.**
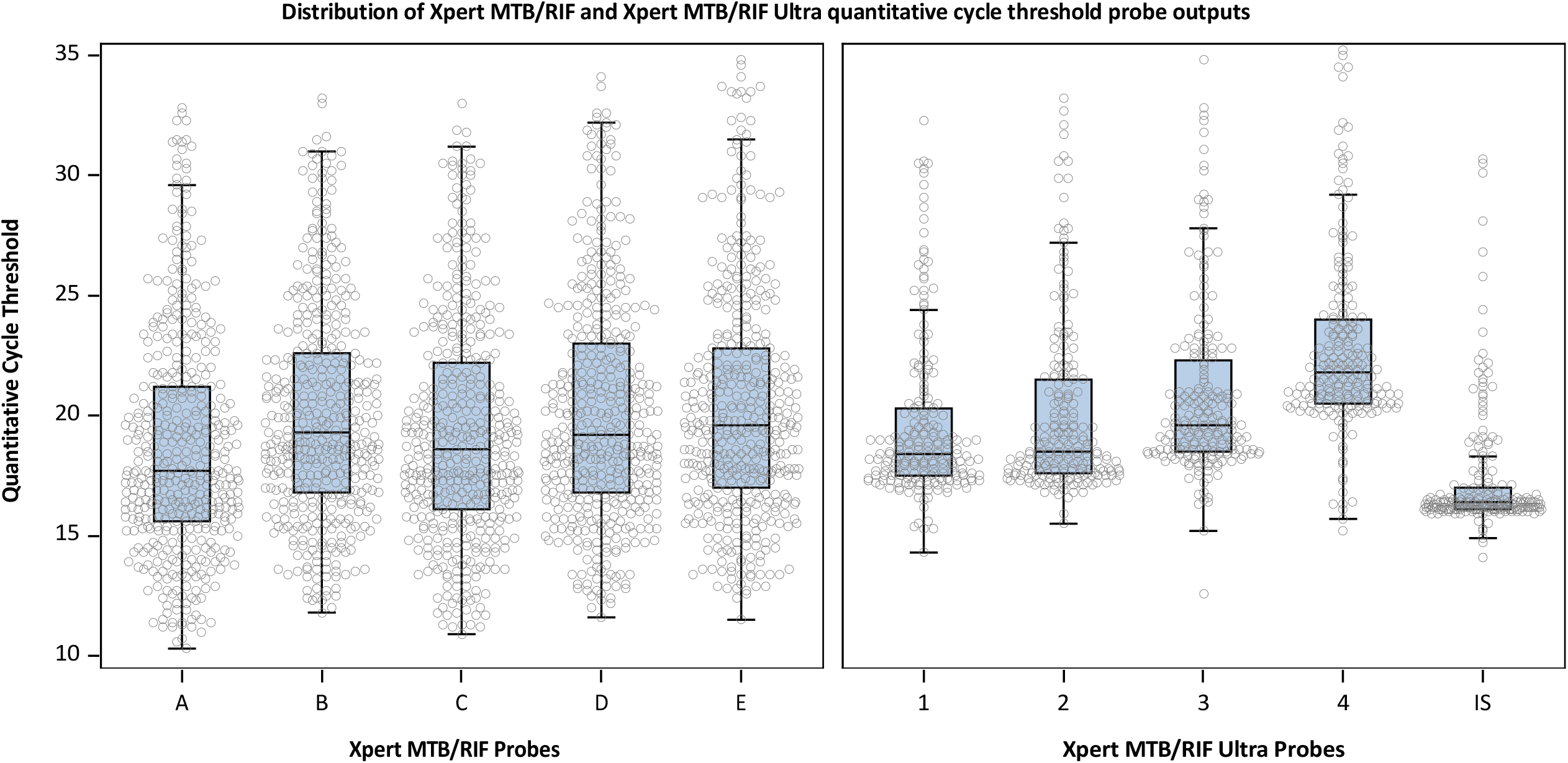
Boxplots showing the distribution of baseline quantitative cycle threshold outputs for each probe of Xpert MTB/RIF (left panel) and Xpert MTB/RIF Ultra (right panel) for the RIFASHORT study population.

There was strong evidence of an association between Xpert quantitative cycle threshold and sputum culture grade, sputum smear grade, and chest X-ray grade (all P<0.001; Figure 2, first row). However, the coefficient of determination varied considerably. Chest X-ray extent of lung disease explained 6.3% of the variability in Xpert quantitative cycle threshold, but had no association with Ultra. Sputum smear grade explained 23.6% and 29.8% of variability in Xpert and Ultra, respectively.

**Figure 2.**
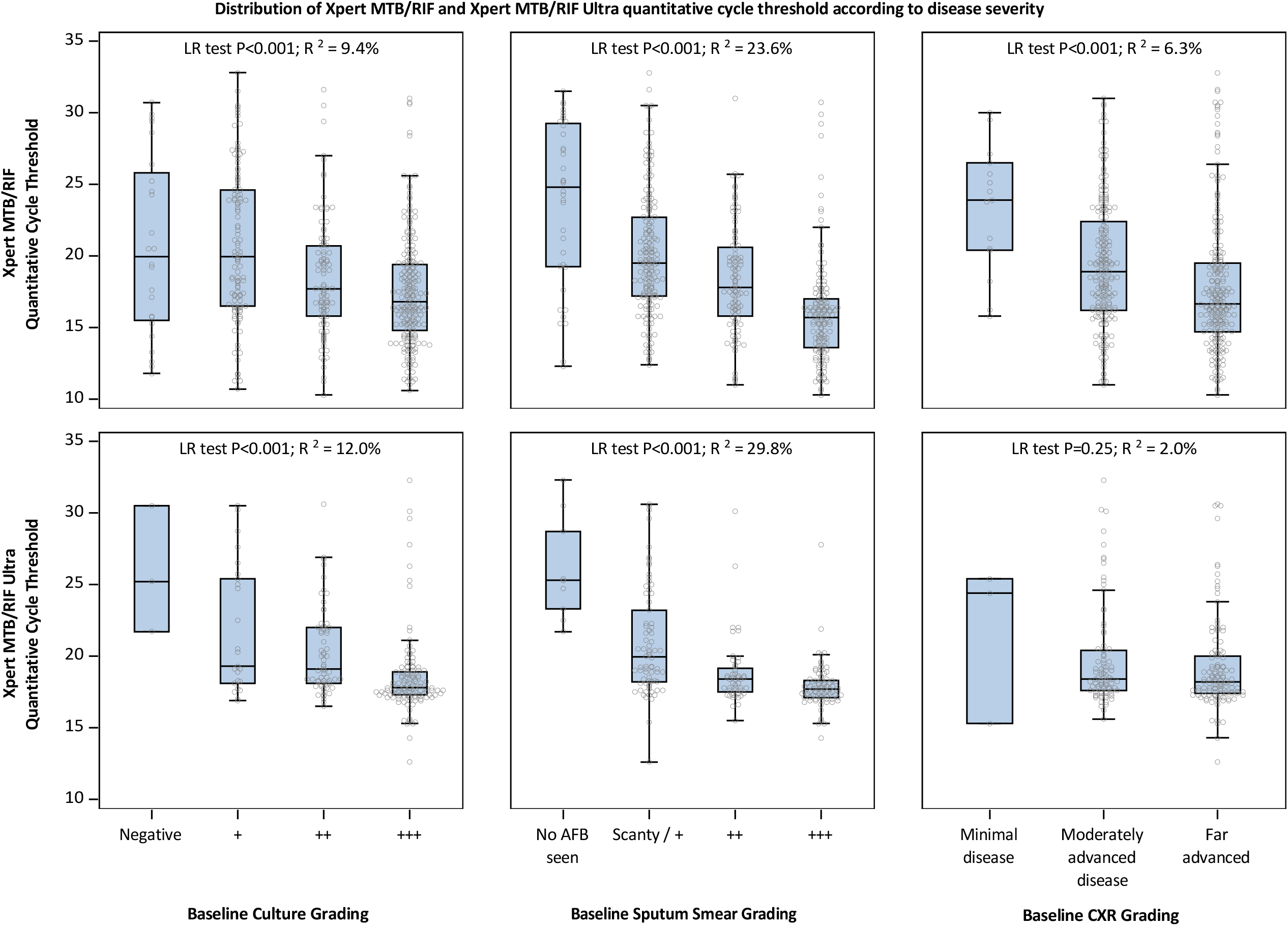
Boxplots showing the distribution of baseline quantitative cycle threshold output for Xpert MTB/RIF (top row) and Xpert MTB/RIF Ultra (bottom row) according to culture grading (first column), sputum smear grading (second column), and chest x-ray grading (third column).

There was greater differentiation in quantitative cycle threshold between the more severe sputum smear grades for Xpert (median: 17.8 ++; 15.7 +++; difference = 2.1) compared with Ultra (median: 18.4 ++; 17.7 +++; difference = 0.7).

### TB phenotype classification

The sensitivity and specificity of each baseline disease severity marker in identifying relapse is shown in Table 2. A high semiquantitative bacterial burden from Xpert or Ultra had a sensitivity of 65.2%, identifying 15/23 relapse cases. However, 160/423 (37.8%) of those who did not relapse also had a high semiquantitative bacterial burden, meaning semiquantitative bacterial burden alone had low specificity (62.2%). Similar patterns were observed for baseline chest X-ray (far advanced: 69.6% sensitivity; 48.1% specificity) and sputum culture (++: 56.5% sensitivity; 50.5% specificity).

**Table 2.**
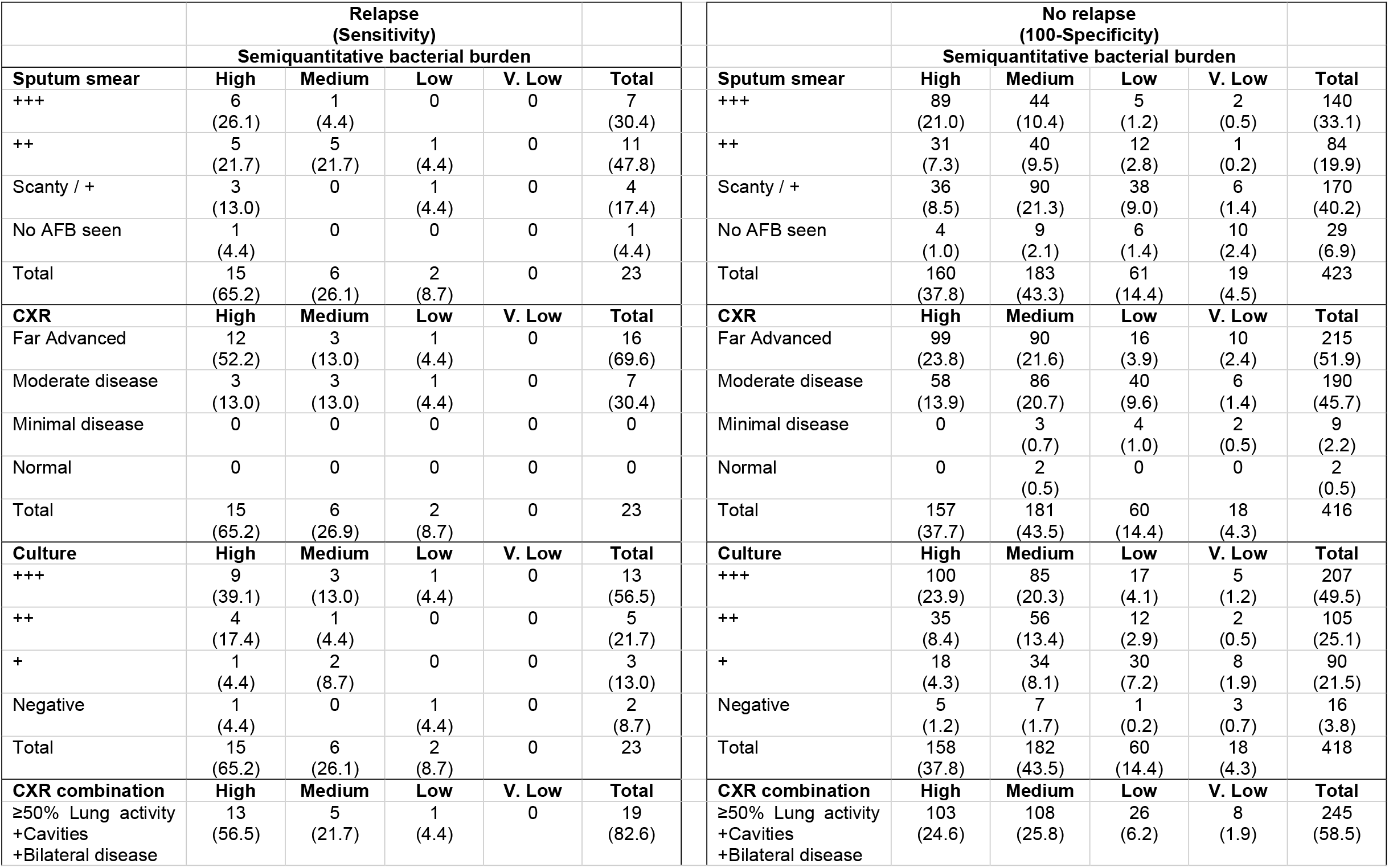
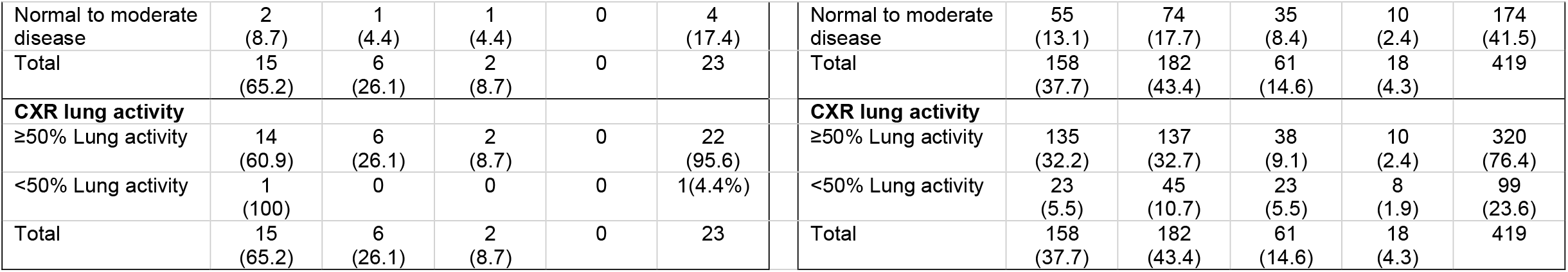
Optimal markers of TB disease severity. The table shows the number of participants who are included in each two-way combination of TB disease burden markers, stratified by TB relapse. In the relapse column, the percentages correspond to the sensitivity of each combination of markers for identifying relapse. In the no relapse column, the percentages correspond to 1-specificity of each combination of markers for no relapse. Total columns show the sensitivity and 100-specificity for individual disease burden markers.

|  | **Relapse**<br>**(Sensitivity)** |  |  |  |  |  | **No relapse**<br>**(100-Specificity)** |  |  |  |  |
| --- | --- | --- | --- | --- | --- | --- | --- | --- | --- | --- | --- |
|  | **Semiquantitative bacterial burden** |  |  |  |  |  | **Semiquantitative bacterial burden** |  |  |  |  |
| **Sputum smear** | **High** | **Medium** | **Low** | **V. Low** | **Total** | **Sputum smear** | **High** | **Medium** | **Low** | **V. Low** | **Total** |
| +++ | 6<br>(26.1) | 1<br>(4.4) | 0 | 0 | 7<br>(30.4) | +++ | 89<br>(21.0) | 44<br>(10.4) | 5<br>(1.2) | 2<br>(0.5) | 140<br>(33.1) |
| ++ | 5<br>(21.7) | 5<br>(21.7) | 1<br>(4.4) | 0 | 11<br>(47.8) | ++ | 31<br>(7.3) | 40<br>(9.5) | 12<br>(2.8) | 1<br>(0.2) | 84<br>(19.9) |
| Scanty / + | 3<br>(13.0) | 0 | 1<br>(4.4) | 0 | 4<br>(17.4) | Scanty / + | 36<br>(8.5) | 90<br>(21.3) | 38<br>(9.0) | 6<br>(1.4) | 170<br>(40.2) |
| No AFB seen | 1<br>(4.4) | 0 | 0 | 0 | 1<br>(4.4) | No AFB seen | 4<br>(1.0) | 9<br>(2.1) | 6<br>(1.4) | 10<br>(2.4) | 29<br>(6.9) |
| Total | 15<br>(65.2) | 6<br>(26.1) | 2<br>(8.7) | 0 | 23 | Total | 160<br>(37.8) | 183<br>(43.3) | 61<br>(14.4) | 19<br>(4.5) | 423 |
| **CXR** | **High** | **Medium** | **Low** | **V. Low** | **Total** | **CXR** | **High** | **Medium** | **Low** | **V. Low** | **Total** |
| Far Advanced | 12<br>(52.2) | 3<br>(13.0) | 1<br>(4.4) | 0 | 16<br>(69.6) | Far Advanced | 99<br>(23.8) | 90<br>(21.6) | 16<br>(3.9) | 10<br>(2.4) | 215<br>(51.9) |
| Moderate disease | 3<br>(13.0) | 3<br>(13.0) | 1<br>(4.4) | 0 | 7<br>(30.4) | Moderate disease | 58<br>(13.9) | 86<br>(20.7) | 40<br>(9.6) | 6<br>(1.4) | 190<br>(45.7) |
| Minimal disease | 0 | 0 | 0 | 0 | 0 | Minimal disease | 0 | 3<br>(0.7) | 4<br>(1.0) | 2<br>(0.5) | 9<br>(2.2) |
| Normal | 0 | 0 | 0 | 0 | 0 | Normal | 0 | 2<br>(0.5) | 0 | 0 | 2<br>(0.5) |
| Total | 15<br>(65.2) | 6<br>(26.9) | 2<br>(8.7) | 0 | 23 | Total | 157<br>(37.7) | 181<br>(43.5) | 60<br>(14.4) | 18<br>(4.3) | 416 |
| **Culture** | **High** | **Medium** | **Low** | **V. Low** | **Total** | **Culture** | **High** | **Medium** | **Low** | **V. Low** | **Total** |
| +++ | 9<br>(39.1) | 3<br>(13.0) | 1<br>(4.4) | 0 | 13<br>(56.5) | +++ | 100<br>(23.9) | 85<br>(20.3) | 17<br>(4.1) | 5<br>(1.2) | 207<br>(49.5) |
| ++ | 4<br>(17.4) | 1<br>(4.4) | 0 | 0 | 5<br>(21.7) | ++ | 35<br>(8.4) | 56<br>(13.4) | 12<br>(2.9) | 2<br>(0.5) | 105<br>(25.1) |
| + | 1<br>(4.4) | 2<br>(8.7) | 0 | 0 | 3<br>(13.0) | + | 18<br>(4.3) | 34<br>(8.1) | 30<br>(7.2) | 8<br>(1.9) | 90<br>(21.5) |
| Negative | 1<br>(4.4) | 0 | 1<br>(4.4) | 0 | 2<br>(8.7) | Negative | 5<br>(1.2) | 7<br>(1.7) | 1<br>(0.2) | 3<br>(0.7) | 16<br>(3.8) |
| Total | 15<br>(65.2) | 6<br>(26.1) | 2<br>(8.7) | 0 | 23 | Total | 158<br>(37.8) | 182<br>(43.5) | 60<br>(14.4) | 18<br>(4.3) | 418 |
| **CXR combination** | **High** | **Medium** | **Low** | **V. Low** | **Total** | **CXR combination** | **High** | **Medium** | **Low** | **V. Low** | **Total** |
| ≥50% Lung activity<br>+Cavities<br>+Bilateral disease | 13<br>(56.5) | 5<br>(21.7) | 1<br>(4.4) | 0 | 19<br>(82.6) | ≥50% Lung activity<br>+Cavities<br>+Bilateral disease | 103<br>(24.6) | 108<br>(25.8) | 26<br>(6.2) | 8<br>(1.9) | 245<br>(58.5) |

|  |  |  |  |  |  |  |  |  |  |  |  |  |
| --- | --- | --- | --- | --- | --- | --- | --- | --- | --- | --- | --- | --- |
| Normal to moderate disease | 2<br>(8.7) | 1<br>(4.4) | 1<br>(4.4) | 0 | 4<br>(17.4) |  | Normal to moderate disease | 55<br>(13.1) | 74<br>(17.7) | 35<br>(8.4) | 10<br>(2.4) | 174<br>(41.5) |
| Total | 15<br>(65.2) | 6<br>(26.1) | 2<br>(8.7) | 0 | 23 |  | Total | 158<br>(37.7) | 182<br>(43.4) | 61<br>(14.6) | 18<br>(4.3) | 419 |
| **CXR lung activity** |  |  |  |  |  |  | **CXR lung activity** |  |  |  |  |  |
| ≥50% Lung activity | 14<br>(60.9) | 6<br>(26.1) | 2<br>(8.7) | 0 | 22<br>(95.6) |  | ≥50% Lung activity | 135<br>(32.2) | 137<br>(32.7) | 38<br>(9.1) | 10<br>(2.4) | 320<br>(76.4) |
| <50% Lung activity | 1<br>(100) | 0 | 0 | 0 | 1(4.4%) |  | <50% Lung activity | 23<br>(5.5) | 45<br>(10.7) | 23<br>(5.5) | 8<br>(1.9) | 99<br>(23.6) |
| Total | 15<br>(65.2) | 6<br>(26.1) | 2<br>(8.7) | 0 | 23 |  | Total | 158<br>(37.7) | 182<br>(43.4) | 61<br>(14.6) | 18<br>(4.3) | 419 |

The highest sensitivity for a single marker was seen for a combined definition of extensive TB disease involvement on chest x-ray. A chest x-ray grading definition requiring ≥50% lung involvement, bilateral disease, and presence of cavities had 8 2.6% sensitivity (capturing 19/23 relapses), however specificity was low (41.5%).

A high Xpert or Ultra semiquantitative bacterial burden in combination with this definition of extensive TB disease involvement on chest x-ray at baseline gave the strongest differentiation between relapse and non-relapse, achieving 56.5% sensitivity (13/23 relapses) and 75.4% specificity; meaning over half of all relapses were identified in one quarter of the study population (26.2% (116/442) meeting the extensive disease classification (Table 2). This combination was therefore selected as the classification criteria for defining limited and extensive TB disease (limited disease: <high bacterial burden or no extensive disease on X-ray; extensive disease: high bacterial burden and extensive disease on X-ray).

### Reanalysis of RIFASHORT trial data by TB phenotype classification

Among those with limited TB disease, the 4-month experimental regimen with 1200mg rifampicin met the protocol defined non-inferiority criterion in both the modified-intention-to-treat (mITT) analysis (adjusted risk difference (aRD): -1.3% (95% confidence interval (CI): -6.7% to 4.0%)) and the per protocol (PP) analysis (aRD: 1.7% (95% CI: -3.8% to 7.1%)) (Figure 3). The full disaggregation of outcome events is available in supplementary Table 1.

**Figure 3.**
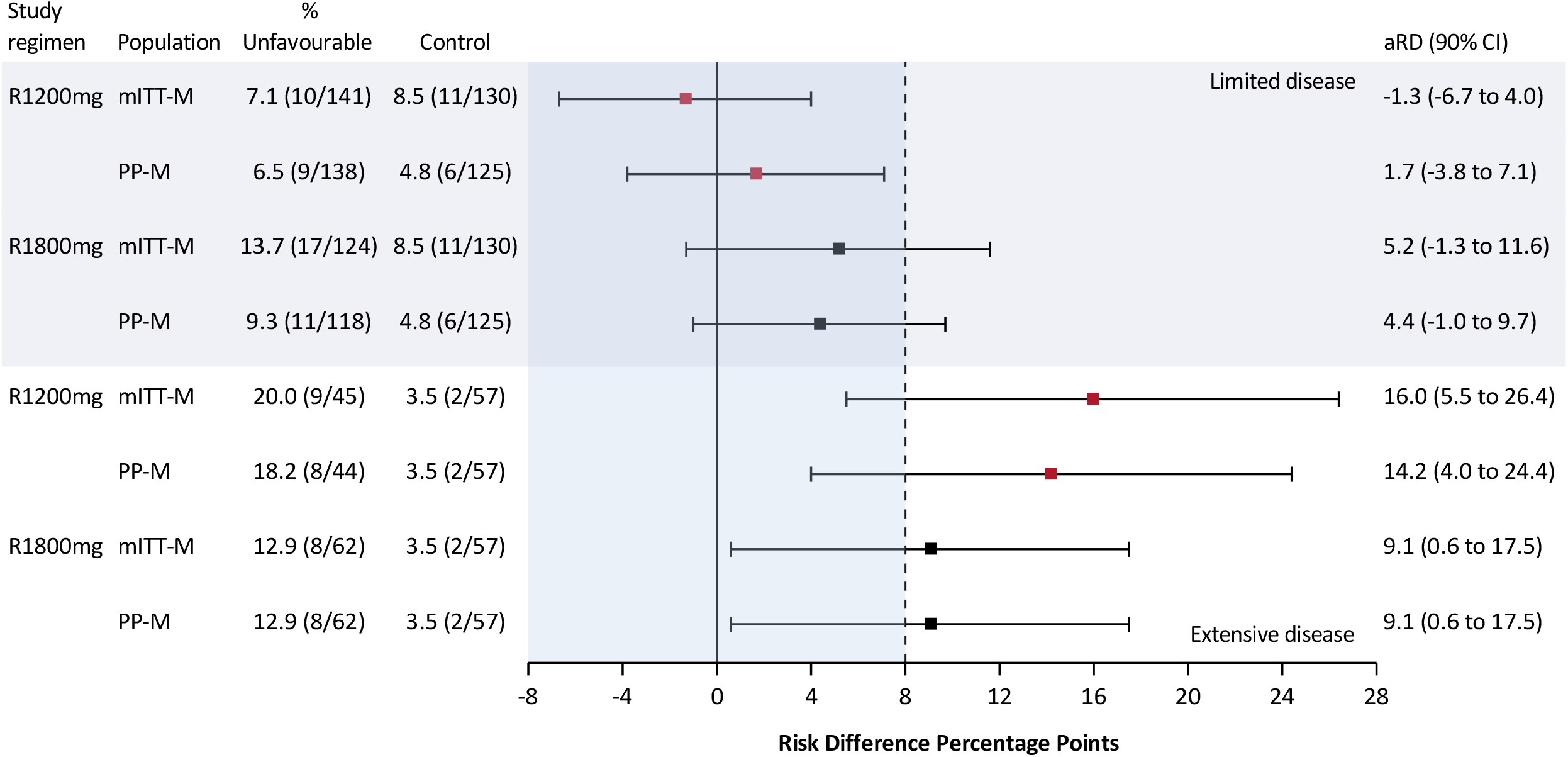
R1200mg: RIFASHORT study regimen using an 1200mg dose of rifampicin; R1800mg: RIFASHORT study regimen using an 1800mg dose of rifampicin; mITT-M: modified intention-to-treat microbiologically positive study population; PP-M: per protocol microbiologically positive study population; aRD: adjusted risk difference; CI: confidence interval.

Among those with extensive TB disease, the 1200mg 4-month experimental regimen performed poorly in comparison to the control. In both mITT (aRD: 16.0% (95% CI: 5.5% to 26.4%)) and PP analysis (aRD: 14.2% (95% CI: 4.0% to 24.4%)) the point estimate of the risk difference to control was greater than 14%.

Similar patterns were observed for the 1800mg 4-month experimental regimen, except it did not achieve non-inferiority among those with limited disease, and performed somewhat better than the 1200mg regimen among those with extensive disease (Figure 3). In the limited disease stratum, the underperformance of the 1800mg rifampicin regimen compared to the 1200mg rifampicin regimen was partially explained by changes in treatment due to adverse events (supplementary Table 1).

In a simplified analysis stratified by semiquantitative bacterial burden only (limited disease: <high; extensive disease: high), 39.2% were classified as having extensive disease, accounting for 15/23 relapses. In the limited disease stratum, the 1200mg rifampicin regimen met the protocol defined non-inferiority criterion in mITT analysis (aRD: 0.0% (95% CI: -6.2% to 6.2%)), but not in PP analysis (aRD: 3.0% (95% CI: -3.2% to 9.2%)) (supplement Figure 1 and Table 2).

## Discussion

A combination of baseline chest X-ray and semiquantitative bacterial burden enabled a binary classification of limited and extensive TB disease. The extensive disease classification accounted for one quarter of RIFASHORT trial participants but more than half of all TB relapses. Reanalysis of the RIFASHORT trial data showed that outcomes in the four-month 1200mg rifampicin arm met the pre-defined criteria for non-inferiority among the three-quarters of the study population with the limited TB disease phenotype. These findings demonstrate the potential for a stratified treatment strategy in determining the duration of TB treatment.

A non-inferior four-month rifapentine-based regimen was reported by Study 31 in 2021,^1^ however, implementation has been hindered by limited access to rifapentine, excessive pill burden, and poor tolerability. A reduction in treatment duration from six to four-months for three-quarters of all drug-susceptible TB patients, using a rifampicin-based regimen would be a major advancement in the field, overcoming challenges faced by the Study 31 regimen and realising cost savings for TB programmes.^12,13^ In the limited disease stratum presented here, the four-month 1200mg rifampicin arm had similar efficacy to the current six-month standard of care, meeting the pre-defined margin of non-inferiority for both mITT and PP analyses. In contrast, the four-month 1800mg rifampicin arm did not demonstrate non-inferiority due to a small number of additional adverse events and treatment withdrawals, suggesting a more favourable tolerability profile for the 1200mg rifampicin arm.

Xpert cycle threshold has been shown to correlate with sputum smear grading,^14^ and other studies have shown an association between Xpert cycle threshold and poor treatment outcomes.^15^ A meta-analysis of three TB treatment duration shortening trials demonstrated the potential for risk stratification tools in the allocation of treatment duration.^16^ However, the risk stratification tool previously proposed required six data inputs, including a sputum culture at two months post-treatment initiation. More recently, Chang et al published a TB disease classification based on the combination of chest X-ray and Xpert quantitative cycle threshold.^17^ Our classification aligns with this, but has one important distinction: using semiquantitative bacterial burden instead of quantitative cycle threshold. The interpretation of semiquantitative bacterial burden is logistically preferable as it is an automatic read out of the machine and requires no interpretation of the quantitative cycle threshold.

The TB phenotype classification developed here requires just two baseline data inputs, Xpert or Ultra semiquantitative bacterial burden and chest X-ray. While chest X-ray is only required in those with a high semiquantitative bacterial burden, chest X-ray will not always be available in the field. Importantly, the 1200mg rifampicin arm had similar efficacy when limited disease was classified based on semiquantitative bacterial burden alone. This means our disease classification may be implemented in settings where chest X-ray is not available, offering potential for individualised TB treatment durations using readily available tools at the point of treatment initiation.

The RIFASHORT trial data are limited in that they exclude people living with HIV, those under 18 years old, and pregnant women. Further, there were few TB relapses in the trial. The limited number of TB relapses restricted our analysis of TB phenotype classification to two-way combinations of disease burden markers. Additional data is required to develop a more sophisticated model accounting for other characteristics. However, the simplicity of the TB phenotype classification may also be seen as a strength regarding ease of implementation. Additional data would also be required to assess the relative sensitivity for TB relapse of Ultra compared to Xpert, given that a higher proportion of participants had a high semiquantitative bacterial burden with Ultra. Better tools for identifying who will TB relapse are still needed. The tools assessed in this study were imprecise individual predictors of TB relapse. There is an important distinction between statistical associations between disease severity markers and treatment outcomes, and individual prognosis, as demonstrated here by strong statistical evidence of associations but with modest sensitivity and specificity of each individual disease severity marker.

In conclusion, the TB phenotype classification derived here successfully identified three-quarters of RIFASHORT trial participants for whom a four-month 1200mg rifampicin regimen was non-inferior to the six-month standard of care. However, this analysis was conducted post-completion of the study; treatment stratification strategies have yet to demonstrate success in prospective randomised controlled trials.^6,7,18^ A four-month rifampicin-based treatment duration for three-quarters of people with TB would be a major advancement in the field and this stratified treatment approach should be assessed in a phase III trial.

## Data Availability

The RIFASHORT trial data are available upon reasonable request via an application to the TB treatment individual patient data platform (TB-IPD). A collaborative initiative to support the generation of reliable evidence on the treatment of TB to inform future TB treatment guidelines. It is supported by the WHO Global TB Programme and maintained by UCL.

https://www.ucl.ac.uk/global-health/research/z-research/tb-ipd-platform

## Acknowledgements

DJG and KF devised the study, DJG performed all statistical analysis and drafted the initial manuscript, JD, PDB, AAW, KG, KL, performed laboratory analysis for the RIFASHORT trial, all other authors contributed to refinements of the analysis and manuscript.

The RIFASHORT trial (https://clinicaltrials.gov/study/NCT02581527) was funded through the MRC/Wellcome Trust/DFID Joint Global Health Trials Scheme (MR/N006127/1) with an additional contribution from the Aga Khan Foundation. This study was completed without funding after completion of the trial.

The authors have no conflicting interests relating to this manuscript.

## The RIFASHORT study team

### St. George’s, University of London

Institute for Infection and Immunity, Division of Clinical Science, St George’s, University of London, Cranmer Terrace, London, SW17 0RE, United Kingdom: Amina Jindani, Thomas Harrison, Philip Butcher, Jasvir Dhillon, Jack Adams, Tulika Munshi, Claire Robb, Nanita Patel, Nandini Shah, Sally Abdelmalik, Sarah Burton, Dr Danni Kirwan, Adam Witney, Katherine Gould, Kenneth Laing.

### The London School of Hygiene and Tropical Medicine

Keppel Street, London, WC1E 7HT, United Kingdom: Professor Katherine Fielding, Dr Daniel Grint.

### University of Leicester

Department of Infection, Immunity and Inflammation, University of Leicester, Leicester: Professor Michael Barer, Jonathan Decker, Galina Mukamolova.

### University of Sussex

Brighton BN1 9PX Medical Research Building, Brighton and Sussex Medical School: Dr Simon Waddell

### Botswana CDC

Botswana, University of Botswana, Gaborone, Botswana: Alyssa Finlay Rosanna Boyd, Dr. Tefera Agizew, Dr Jose Gaby Tshikuka, Dr John T Tlhakanelo, Anikie Mathoma, Unami Mathebula, Joyce Basotli, Norah Dudzai Mawoko, Gagoope Mowaneg, Evelyn Dintwa, Maitumelo Masole, Malebogo Masono

### Uganda

MSF-Epicentre, PO Box 1956, Mbarara, Uganda: Dr Daniel Atwine, Dr Adolf Byamukama, Ivan Mugisha Taremwa, Patrick Orikiriza, Deborah Nanjebe K.T., Dan Nyehangane, Dr Keneth Kananura, Dr Rachel Kyohairwe, Susan Lagoose Akatuhebwa, Dinnah Kansime Lydia, Amelia Arimpa, Yvonne Sheila Mbabazi, Derek Kamulegeya, Eva Natukunda, Brenda Asiimwe, Emmanuel Mucunguzi, Rinah Arinaitwe, Dianah Birungi, Claire Nimusima, Justus Ashaba

### TransVIHMI – University of Montpellier – INSERM

U1175 & Epicentre, Kampala, Uganda: Dr. Maryline Bonnet, Institut de Recherche pour le Développement, UMI 233

### Guinea

Service de Pneumo-Phtisiologie, Centre Hospitalier Universitaire Ignace Deen, B.P: 634, Conakry, République De Guinée: Prof Lansana Mady Camara, Prof Oumou Younoussa Bah, Dr Boubacar Bah, Fulgence Nzabintwali, Dr Diallo Alhassane Djeima, Mme Nene Mamata Mme, Sow Maimouna, Mme Aissatou Bah, Moussa Conde, Kindy Sadjo Bah, Falilou Bah, Sylla Karamoko, Abdourahmane Barry, Mme Kadiatou Balde, Abdourahmane Bah Harouna, Barry Mamadou, Saliou Barry, Alpha Saliou Barry

### Lima

Hospital Dos de Mayo and Universidad Nacional Mayor de San Marcos (UNMSM), Lima, Peru: Dr. Eduardo Ticona Chávez, Dra. Luz Huaroto, Dr. Victor Manuel Chávez Pérez, Dr. Juan Genaro Sosa Páucar, Dr. César Eduardo Ticona Huaroto Dra. Paola Rondan Raquel Mugruza Pineda, Wendy Shulay Guevara Condorhuamán, Gloria Espinoza Vergaray, Verónica Carmona Flores, Julio Tanta Vergaray, Luis Vizuña Lorenzo, Maria Elena Vilca Valverde, María Isabel López Moreno, Wilfredo Vargas Onofre, Karín Canchari Salazar, Mónica Basauri Infantes, Ynocente Raúl Joaquín Vásquez, Jessica Chávez Echevarría, Rossana Pacheco Aguilar, Gaby Evelyn Alvarez Mantari, Miriam Quispe Quispe, Ismael Armando Collantes Diaz, José Julio Zúñiga Cruz. University of New Mexico, Albuquerque, USA: Dr. Marcos Burgos

### Nepal

GENETUP, PO Box 1494, Kalimati, Kathmandu, Nepal: Dr Bhabana Shrestha, Dr. Neko Mitra Shrestha, Sanu Tandukar, Bhagwan Maharjan, Bijesh Tandukar, Bijendra Bhakta Raya, Matina Maharjan, Samriddhi Karki, Meera Shrestha.

### Pakistan

Department of Pathology/Microbiology, Aga Khan University, Karachi. Pakistan: Dr Saeed Hamid, Dr Bushra Jamil, Dilshad Begum, Dr Imran Ahmed, Professor Rumina Hasan, Dr Irfan Muhammad, Ammara Muzammil, Dr Paras Shahzad, Dr Anum Adil, Dr Hira Jawed. Shaukat Khanum Memorial & Cancer Research Centre, Lahore, Pakistan: Dr. Faisal Sultan, Dr. Mariam Hassan, Dr. Iqra Masood, Dr. Summiya Nizamuddin.

## Notes

### Competing Interest Statement

The authors have declared no competing interest.

### Clinical Trial

NCT02581527

### Author Declarations

The RIFASHORT trial was approved by the London School of Hygiene and Tropical Medicine Research Ethics Committee, as well as institutional and national ethics and regulatory authorities representing all participating sites and countries.

