## Supplementary material for "Xpert MTB/RIF^®^ cycle threshold as a marker of TB disease severity; Implications for TB treatment stratification"

### 1. RIFASHORT trial chest X-ray case report form (CRF)

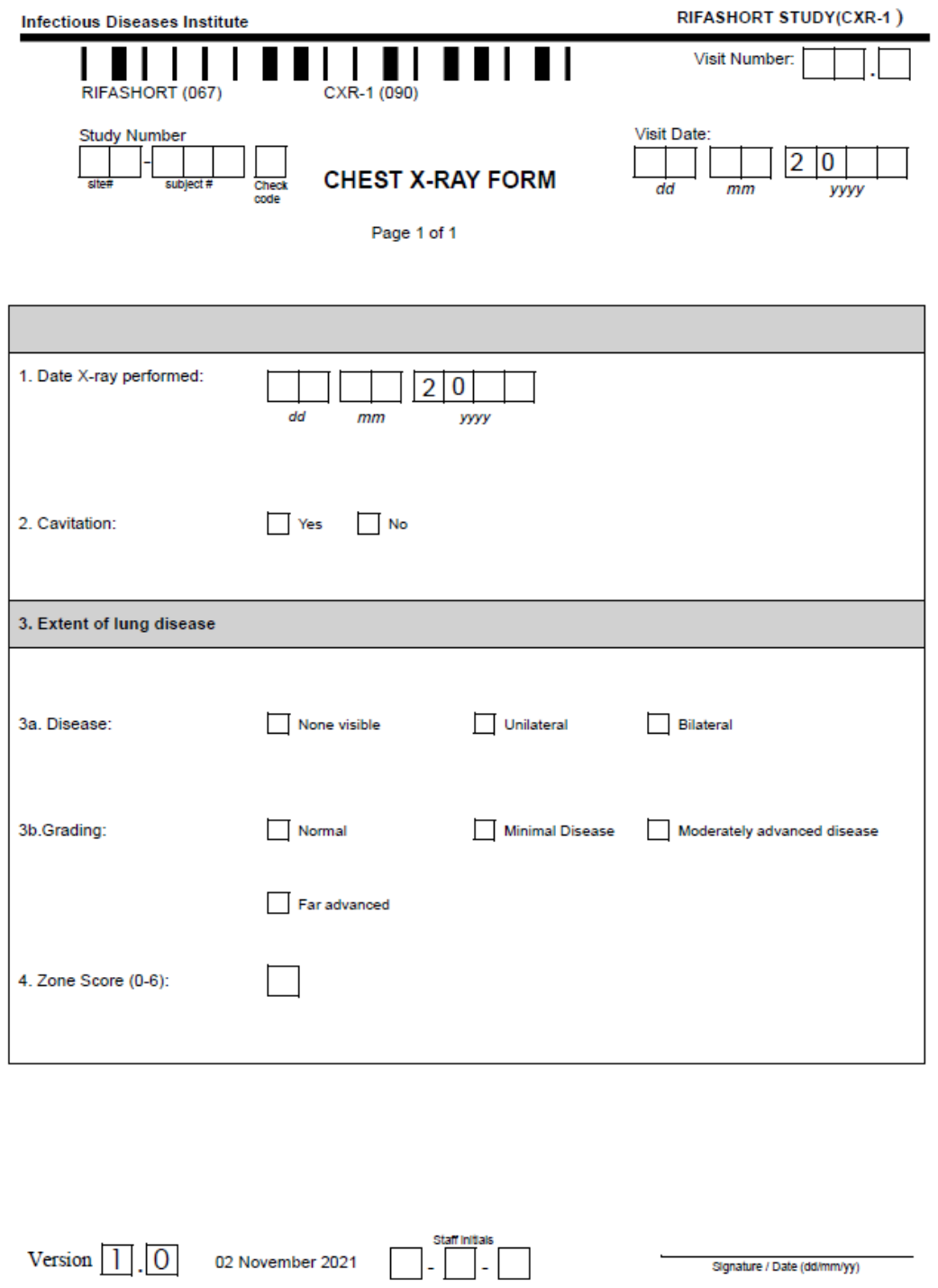

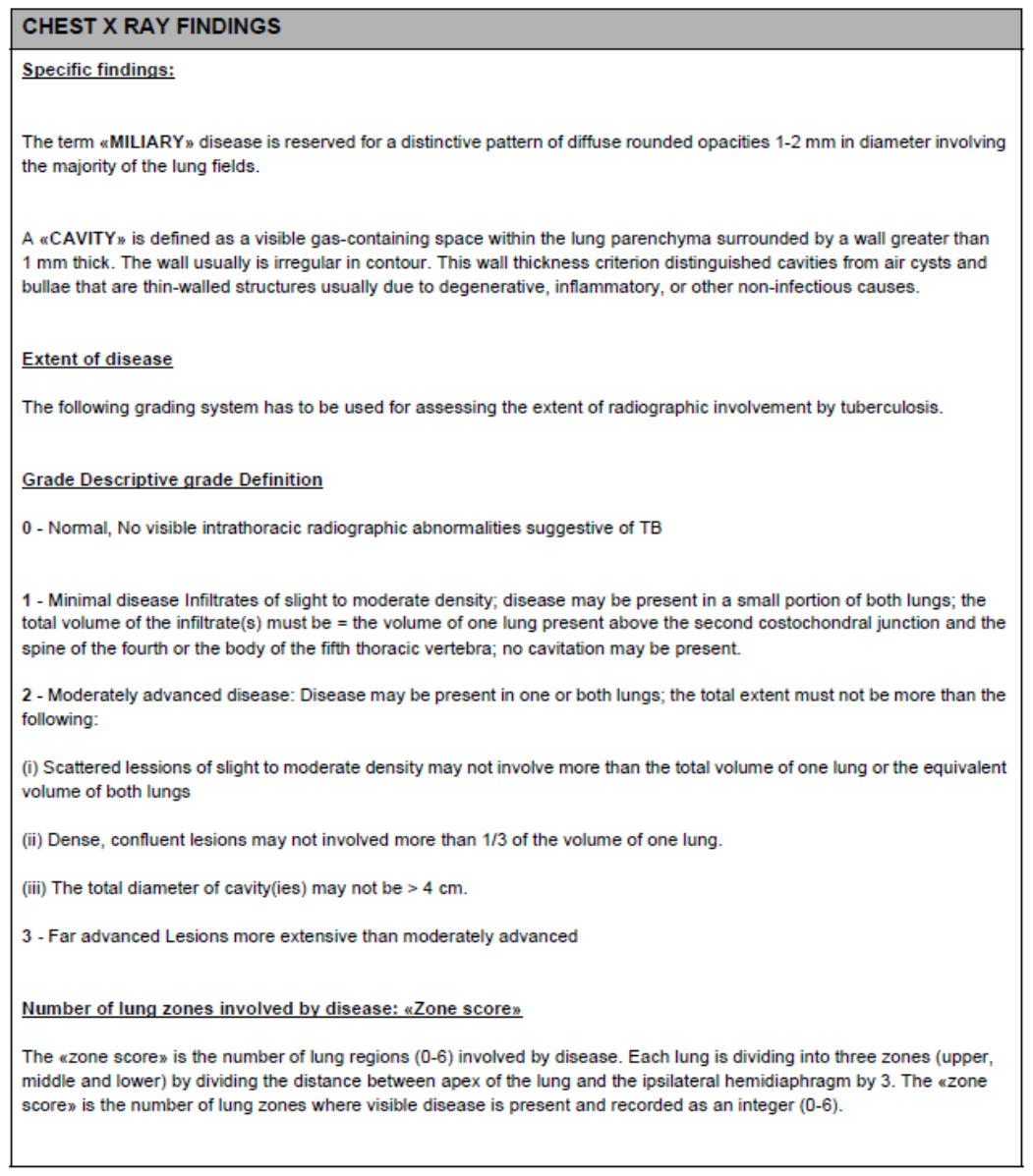

### 2. Excerpt on analysis populations from the RIFASHORT trial statistical analysis plan (SAP)

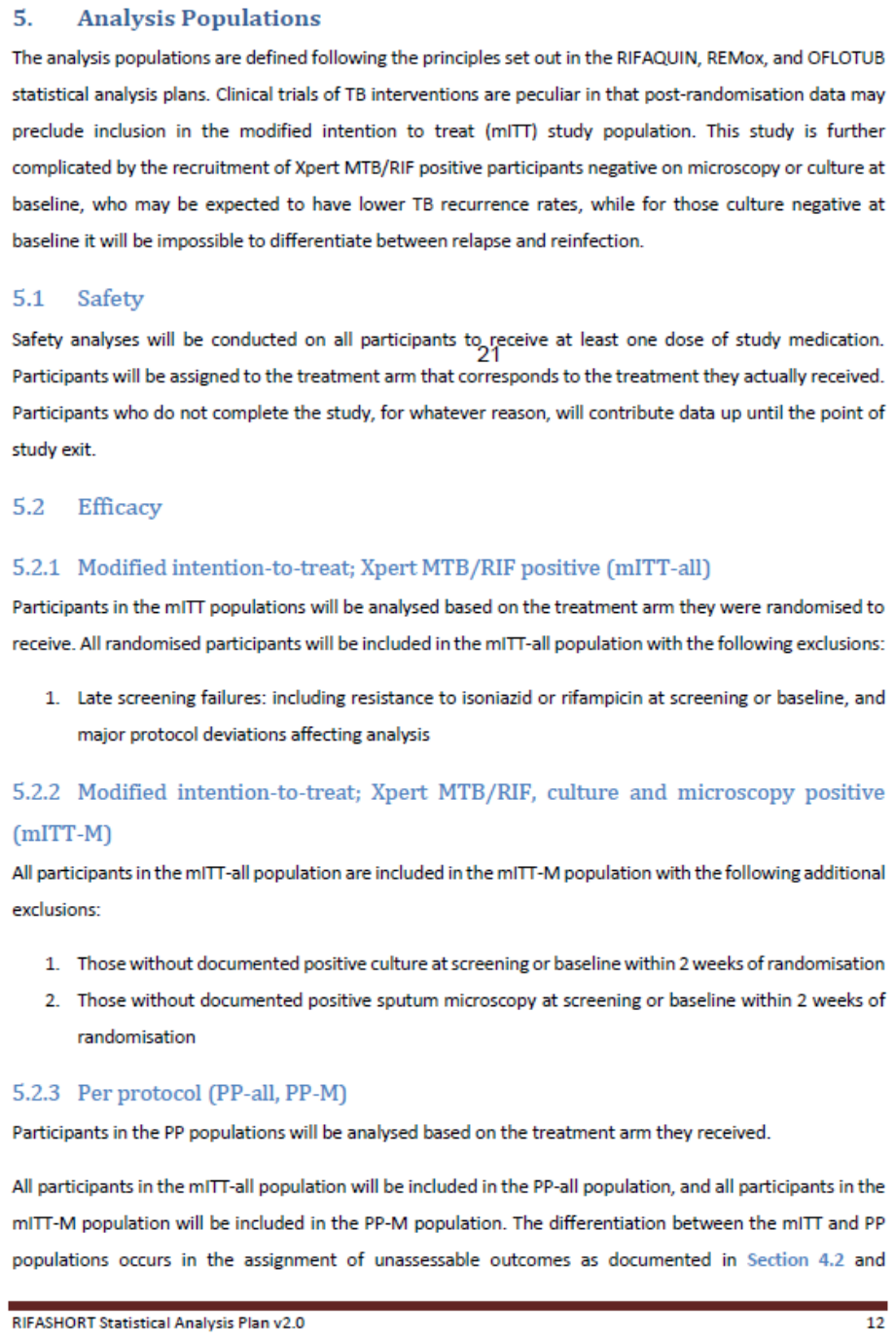

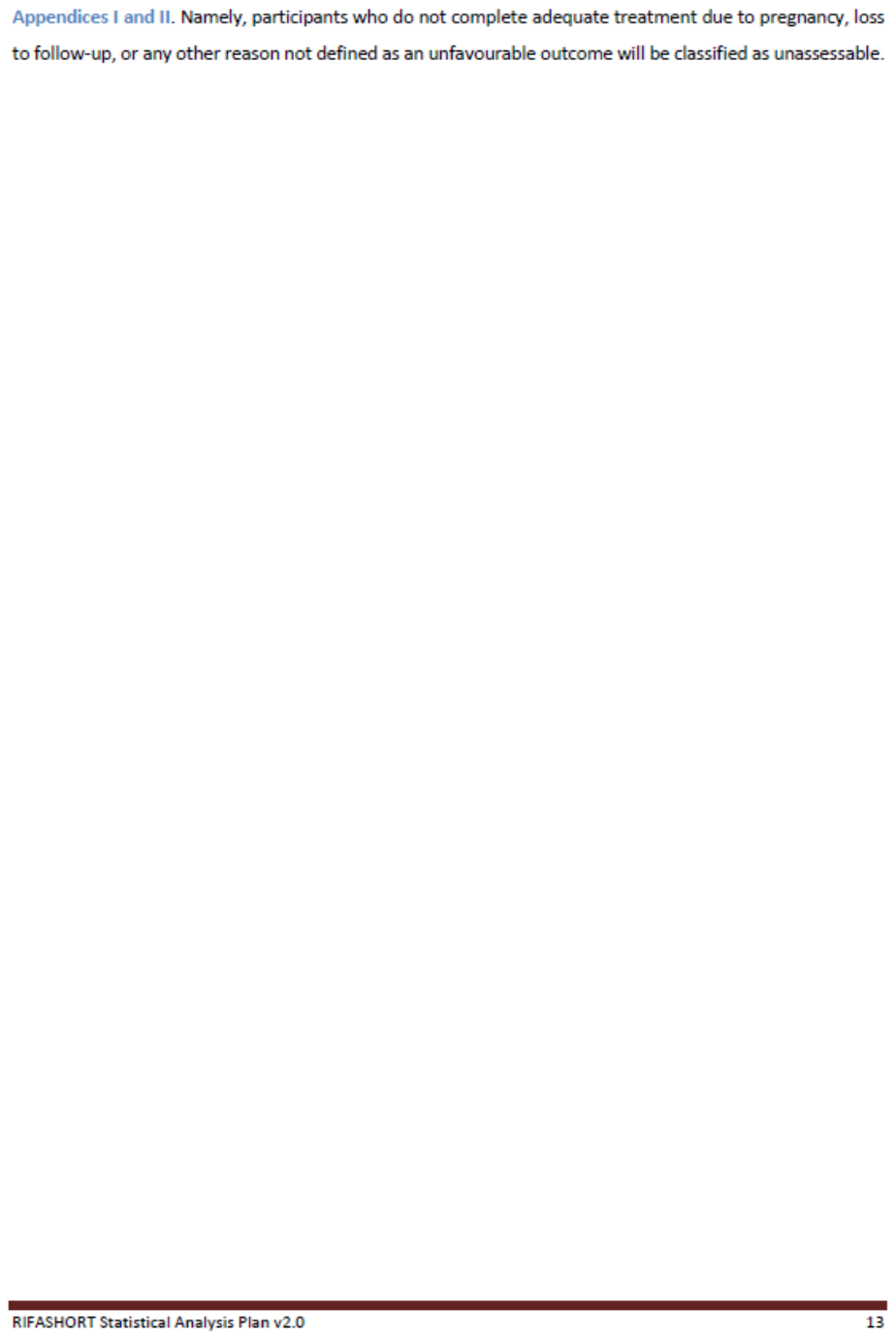

### Table 1 RIFASHORT trial outcome disaggregation according to limited and extensive disease

**Limited disease outcomes:** defined by absence of CXR (≥50% lung involvement, bilateral disease, and cavitation) and <high semiquantitative CT

| **mITT-M** analysis outcomes | Control (N=130) | Study regimen 1 (N=141) | Study regimen 2 (N=124) |
| --- | --- | --- | --- |
| Favourable |  |  |  |
| Participants with outcome - n (%) | 119 (91.5) | 131 (92.9) | 107 (86.3) |
| Unfavourable |  |  |  |
| Participants with outcome - n (%) | 11 (8.5) | 10 (7.1) | 17 (13.7) |
| Adjusted risk difference to control (95% CI) |  |  |  |
| Death during the treatment phase | 2 (1.5) | 3 (2.1) | 0 |
| Post-treatment death, TB a plausible cause | 0 | 1 (0.7) | 0 |
| Lost to follow-up during the treatment phase | 2 (1.5) | 0 | 1 (0.8) |
| Withdrew from the trial during the treatment phase | 3 (2.3) | 1 (0.7) | 5 (4.0) |
| Change in treatment due to adverse event | 1 (0.8) | 1 (0.7) | 5 (4.0) |
| Two consecutive positive cultures after completing treatment | 1 (0.8) | 3 (2.1) | 3 (2.4) |
| Retreated for TB due to clinical signs and symptoms without two consecutive positive cultures | 2 (1.5) | 1 (0.7) | 3 (2.4) |
| Exclusions |  |  |  |
| Late exclusion - Baseline INH resistance | 8 | 7 | 8 |
| Late exclusion - Culture negative | 0 | 0 | 2 |
| Late exclusion - <18 years old | 0 | 1 | 0 |
| Late exclusion - Medical history that should have excluded randomisation | 0 | 0 | 2 |
| Late exclusion - Extrapulmonary TB at baseline on assessment | 1 | 1 | 0 |
| Unassessable - Post-treatment death deemed unrelated to TB or treatment | 0 | 1 | 0 |
| Unassessable - Post-treatment LTFU when culture negative | 1 | 3 | 5 |
| Unassessable - Evidence of exogenous TB reinfection | 0 | 0 | 2 |
| Unassessable - Withdrawal during the treatment phase when culture negative | 1 | 0 | 0 |
| Unassessable - Post-treatment withdrawal when culture negative | 0 | 1 | 0 |
| Exclusion - No evidence of culture or smear positivity at baseline | 18 | 18 | 14 |

| **PP-M** analysis outcomes | Control (N=125) | Study regimen 1 (N=138) | Study regimen 2 (N=118) |
| --- | --- | --- | --- |
| Favourable |  |  |  |
| Participants with outcome - n (%) | 119 (95.2) | 129 (93.5) | 107 (90.7) |
| Unfavourable |  |  |  |
| Participants with outcome - n (%) | 6 (4.8) | 9 (6.5) | 11 (9.3) |
| Adjusted risk difference to control (95% CI) |  |  |  |
| Death during the treatment phase | 2 (1.6) | 3 (2.2) | 0 |
| Post-treatment death, TB a plausible cause | 0 | 1 (0.7) | 0 |
| Change in treatment due to adverse event | 1 (0.8) | 1 (0.7) | 5 (4.2) |
| Two consecutive positive cultures after completing treatment | 1 (0.8) | 3 (2.2) | 3 (2.5) |
| Retreated for TB due to clinical signs and symptoms without two consecutive positive cultures | 2 (1.6) | 1 (0.7) | 3 (2.5) |
| Exclusions |  |  |  |
| Late exclusion - Baseline INH resistance | 8 | 7 | 8 |
| Late exclusion - Culture negative | 0 | 0 | 2 |
| Late exclusion - <18 years old | 0 | 1 | 0 |
| Late exclusion - Medical history that should have excluded randomisation | 0 | 0 | 2 |
| Late exclusion - Extrapulmonary TB at baseline on assessment | 1 | 1 | 0 |
| Unassessable - Post-treatment death deemed unrelated to TB or treatment | 0 | 1 | 0 |
| Unassessable - Treatment phase LTFU | 2 | 0 | 1 |
| Unassessable - Post-treatment LTFU when culture negative | 1 | 3 | 5 |
| Unassessable - Evidence of exogenous TB reinfection | 0 | 0 | 2 |
| Unassessable - Withdrawal during the treatment phase | 3 | 1 | 5 |
| Unassessable - Withdrawal during the treatment phase when culture negative | 1 | 0 | 0 |
| Unassessable - Post-treatment withdrawal when culture negative | 0 | 1 | 0 |
| Unassessable - Switched to control regimen due to pregnancy | 0 | 2 | 0 |
| Exclusion - No evidence of culture or smear positivity at baseline | 18 | 18 | 14 |

**Extensive disease outcomes:** defined by CXR (≥50% lung involvement, bilateral disease, and cavitation) and high semiquantitative CT

| **mITT-M** analysis outcomes | Control (N=57) | Study regimen 1 (N=45) | Study regimen 2 (N=62) |
| --- | --- | --- | --- |
| Favourable |  |  |  |
| Participants with outcome - n (%) | 55 (96.5) | 36 (80.0) | 54 (87.1) |
| Unfavourable |  |  |  |
| Participants with outcome - n (%) | 2 (3.5) | 9 (20.0) | 8 (12.9) |
| Adjusted risk difference to control (95% CI) |  |  |  |
| Death during the treatment phase | 1 (1.8) | 1 (2.2) | 0 |
| Withdrew from the trial during the treatment phase | 0 | 1 (2.2) | 0 |
| Change in treatment due to adverse event | 0 | 1 (2.2) | 2 (3.2) |
| Two consecutive positive cultures after completing treatment | 1 (1.8) | 6 (13.3) | 6 (9.7) |
| Exclusions |  |  |  |
| Late exclusion - Baseline INH resistance | 3 | 2 | 1 |
| Unassessable - Post-treatment death deemed unrelated to TB or treatment | 2 | 0 | 2 |
| Unassessable - Evidence of exogenous TB reinfection | 0 | 1 | 0 |
| Exclusion - No evidence of culture or smear positivity at baseline | 1 | 2 | 1 |

| **PP-M** analysis outcomes | Control (N=57) | Study regimen 1 (N=44) | Study regimen 2 (N=62) |
| --- | --- | --- | --- |
| Favourable |  |  |  |
| Participants with outcome - n (%) | 55 (96.5) | 36 (81.8) | 54 (87.1) |
| Unfavourable |  |  |  |
| Participants with outcome - n (%) | 2 (3.5) | 8 (18.2) | 8 (12.9) |
| Adjusted risk difference to control (95% CI) |  |  |  |
| Death during the treatment phase | 1 (1.8) | 1 (2.3) | 0 |
| Change in treatment due to adverse event | 0 | 1 (2.3) | 2 (3.2) |
| Two consecutive positive cultures after completing treatment | 1 (1.8) | 6 (13.6) | 6 (9.7) |
| Exclusions |  |  |  |
| Late exclusion - Baseline INH resistance | 3 | 2 | 1 |
| Unassessable - Post-treatment death deemed unrelated to TB or treatment | 2 | 0 | 2 |
| Unassessable - Evidence of exogenous TB reinfection | 0 | 1 | 0 |
| Unassessable - Withdrawal during the treatment phase | 0 | 1 | 0 |
| Exclusion - No evidence of culture or smear positivity at baseline | 1 | 2 | 1 |

### Figure 1 RIFASHORT reanalysis with stratification by semiquantitative bacterial burden only

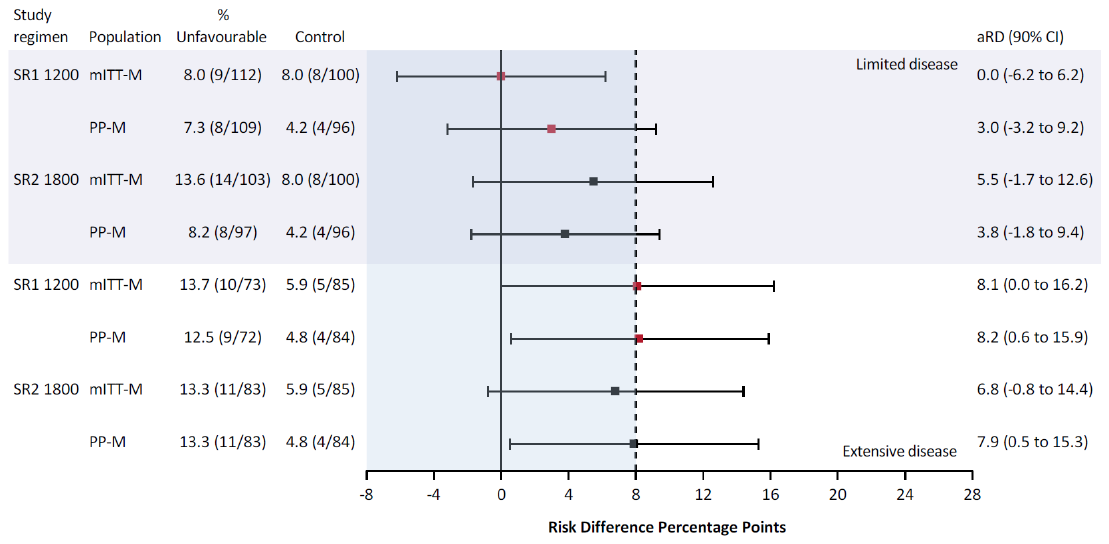

### Table 2 RIFASHORT trial outcome disaggregation according to limited and extensive disease defined by semiquantitative bacterial burden only

**Limited disease outcomes –** defined by CT <high only

| **mITT-M** analysis outcomes | Control (N=100) | Study regimen 1 (N=112) | Study regimen 2 (N=103) |
| --- | --- | --- | --- |
| Favourable |  |  |  |
| Participants with outcome - n (%) | 92 (92.0) | 103 (92.0) | 89 (86.4) |
| Unfavourable |  |  |  |
| Participants with outcome - n (%) | 8 (8.0) | 9 (8.0) | 14 (13.6) |
| Adjusted risk difference to control (95% CI) |  |  |  |
| Death during the treatment phase | 1 (1.0) | 3 (2.7) | 0 |
| Post-treatment death, TB a plausible cause | 0 | 1 (0.9) | 0 |
| Lost to follow-up during the treatment phase | 2 (2.0) | 0 | 1 (1.0) |
| Withdrew from the trial during the treatment phase | 2 (2.0) | 1 (0.9) | 5 (4.9) |
| Change in treatment due to adverse event | 1 (1.0) | 0 | 4 (3.9) |
| Two consecutive positive cultures after completing treatment | 1 (1.0) | 3 (2.7) | 2 (1.9) |
| Retreated for TB due to clinical signs and symptoms without two consecutive positive cultures | 1 (1.0) | 1 (0.9) | 2 (1.9) |
| Exclusions |  |  |  |
| Late exclusion - Baseline INH resistance | 5 | 7 | 7 |
| Late exclusion - Culture negative | 0 | 0 | 1 |
| Late exclusion - Medical history that should have excluded randomisation | 0 | 0 | 2 |
| Late exclusion - Extrapulmonary TB at baseline on assessment | 1 | 1 | 0 |
| Unassessable - Post-treatment death deemed unrelated to TB or treatment | 0 | 1 | 0 |
| Unassessable - Post-treatment LTFU when culture negative | 1 | 2 | 4 |
| Unassessable - Evidence of exogenous TB reinfection | 0 | 0 | 2 |
| Unassessable - Withdrawal during the treatment phase when culture negative | 1 | 0 | 0 |
| Unassessable - Post-treatment withdrawal when culture negative | 0 | 1 | 0 |
| Exclusion - No evidence of culture or smear positivity at baseline | 16 | 15 | 11 |

| **PP-M** analysis outcomes | Control (N=96) | Study regimen 1 (N=109) | Study regimen 2 (N=97) |
| --- | --- | --- | --- |
| Favourable |  |  |  |
| Participants with outcome - n (%) | 92 (95.8) | 101 (92.7) | 89 (91.8) |
| Unfavourable |  |  |  |
| Participants with outcome - n (%) | 4 (4.2) | 8 (7.3) | 8 (8.2) |
| Adjusted risk difference to control (95% CI) |  |  |  |
| Death during the treatment phase | 1 (1.0) | 3 (2.8) | 0 |
| Post-treatment death, TB a plausible cause | 0 | 1 (0.9) | 0 |
| Change in treatment due to adverse event | 1 (1.0) | 0 | 4 (4.1) |
| Two consecutive positive cultures after completing treatment | 1 (1.0) | 3 (2.8) | 2 (2.1) |
| Retreated for TB due to clinical signs and symptoms without two consecutive positive cultures | 1 (1.0) | 1 (0.9) | 2 (2.1) |
| Exclusions |  |  |  |
| Late exclusion - Baseline INH resistance | 5 | 7 | 7 |
| Late exclusion - Culture negative | 0 | 0 | 1 |
| Late exclusion - Medical history that should have excluded randomisation | 0 | 0 | 2 |
| Late exclusion - Extrapulmonary TB at baseline on assessment | 1 | 1 | 0 |
| Unassessable - Post-treatment death deemed unrelated to TB or treatment | 0 | 1 | 0 |
| Unassessable - Treatment phase LTFU | 2 | 0 | 1 |
| Unassessable - Post-treatment LTFU when culture negative | 1 | 2 | 4 |
| Unassessable - Evidence of exogenous TB reinfection | 0 | 0 | 2 |
| Unassessable - Withdrawal during the treatment phase | 2 | 1 | 5 |
| Unassessable - Withdrawal during the treatment phase when culture negative | 1 | 0 | 0 |
| Unassessable - Post-treatment withdrawal when culture negative | 0 | 1 | 0 |
| Unassessable - Switched to control regimen due to pregnancy | 0 | 2 | 0 |
| Exclusion - No evidence of culture or smear positivity at baseline | 16 | 15 | 11 |

**Extensive disease outcomes:** defined by CT high only

| **mITT-M** analysis outcomes | Control (N=85) | Study regimen 1 (N=73) | Study regimen 2 (N=83) |
| --- | --- | --- | --- |
| Favourable |  |  |  |
| Participants with outcome - n (%) | 80 (94.1) | 63 (86.3) | 72 (86.7) |
| Unfavourable |  |  |  |
| Participants with outcome - n (%) | 5 (5.9) | 10 (13.7) | 11 (13.3) |
| Adjusted risk difference to control (95% CI) |  |  |  |
| Death during the treatment phase | 2 (2.4) | 1 (1.4) | 0 |
| Withdrew from the trial during the treatment phase | 1 (1.2) | 1 (1.4) | 0 |
| Change in treatment due to adverse event | 0 | 2 (2.7) | 3 (3.6) |
| Two consecutive positive cultures after completing treatment | 1 (1.2) | 6 (8.2) | 7 (8.4) |
| Retreated for TB due to clinical signs and symptoms without two consecutive positive cultures | 1 (1.2) | 0 | 1 (1.2) |
| Exclusions |  |  |  |
| Late exclusion - Baseline INH resistance | 6 | 2 | 2 |
| Late exclusion - Culture negative | 0 | 0 | 1 |
| Late exclusion - <18 years old | 0 | 1 | 0 |
| Unassessable - Post-treatment death deemed unrelated to TB or treatment | 2 | 0 | 2 |
| Unassessable - Post-treatment LTFU when culture negative | 0 | 1 | 1 |
| Unassessable - Evidence of exogenous TB reinfection | 0 | 1 | 0 |
| Exclusion - No evidence of culture or smear positivity at baseline | 2 | 5 | 3 |

| **PP-M** analysis outcomes | Control (N=84) | Study regimen 1 (N=72) | Study regimen 2 (N=83) |
| --- | --- | --- | --- |
| Favourable |  |  |  |
| Participants with outcome - n (%) | 80 (95.2) | 63 (87.5) | 72 (86.7) |
| Unfavourable |  |  |  |
| Participants with outcome - n (%) | 4 (4.8) | 9 (12.5) | 11 (13.3) |
| Adjusted risk difference to control (95% CI) |  |  |  |
| Death during the treatment phase | 2 (2.4) | 1 (1.4) | 0 |
| Change in treatment due to adverse event | 0 | 2 (2.8) | 3 (3.6) |
| Two consecutive positive cultures after completing treatment | 1 (1.2) | 6 (8.3) | 7 (8.4) |
| Retreated for TB due to clinical signs and symptoms without two consecutive positive cultures | 1 (1.2) | 0 | 1 (1.2) |
| Exclusions |  |  |  |
| Late exclusion - Baseline INH resistance | 6 | 2 | 2 |
| Late exclusion - Culture negative | 0 | 0 | 1 |
| Late exclusion - <18 years old | 0 | 1 | 0 |
| Unassessable - Post-treatment death deemed unrelated to TB or treatment | 2 | 0 | 2 |
| Unassessable - Post-treatment LTFU when culture negative | 0 | 1 | 1 |
| Unassessable - Evidence of exogenous TB reinfection | 0 | 1 | 0 |
| Unassessable - Withdrawal during the treatment phase | 1 | 1 | 0 |
| Exclusion - No evidence of culture or smear positivity at baseline | 2 | 5 | 3 |
